# Brain and systemic inflammation in *de novo* Parkinson’s disease

**DOI:** 10.1101/2022.09.21.22280194

**Authors:** Talene A. Yacoubian, Yu-Hua Dean Fang, Adam Gerstenecker, Amy Amara, Natividad Stover, Lauren Ruffrage, Christopher Collette, Richard Kennedy, Yue Zhang, Huixian Hong, Hongwei Qin, Jonathan McConathy, Etty N. Benveniste, David G. Standaert

**Author notes:** **Corresponding Author:** Talene A. Yacoubian, MD, PhD, 1719 Sixth Avenue South, Civitan International Research Building 510A, Birmingham, AL 35294. **Funding source:** NINDS P50NS108675.

## Abstract

**Objective:** To assess the presence of brain and systemic inflammation in subjects newly diagnosed with Parkinson’s disease (PD).

**Background:** Evidence for a pathophysiologic role of inflammation in PD is growing. However, several key gaps remain as to the role of inflammation in PD, including the extent of immune activation at early stages, potential effects of PD treatments on inflammation, and whether pro-inflammatory signals are associated with clinical features and/or predict more rapid progression.

**Methods:** We enrolled subjects with *de novo* PD (n=58) and age-matched controls (n=62). Subjects underwent clinical assessments, including the Movement Disorder Society-United Parkinson’s Disease rating scale (MDS-UPDRS). Comprehensive cognitive assessment meeting MDS Level II criteria for mild cognitive impairment (MCI) testing was performed. Blood was obtained for flow cytometry and cytokine/chemokine analyses. Subjects underwent imaging with ^18^F-DPA-714, a translocator protein 18kd (TSPO) ligand, and lumbar puncture if eligible and consented.

**Results:** Baseline demographics and medical history were comparable between groups. PD subjects showed significant differences in University of Pennsylvania Smell Identification Test, Schwab and England Activities of Daily Living, Scales for Outcomes in PD autonomic dysfunction, and MDS-UPDRS scores. Cognitive testing demonstrated significant differences in cognitive composite, executive function, and visuospatial domain scores at baseline. PET imaging showed increased ^18^F-DPA-714 signal in PD subjects. ^18^F-DPA-714 signal correlated with several cognitive measures and some chemokines.

**Conclusions:** ^18^F-DPA-714 imaging demonstrated increased central inflammation in *de novo* PD subjects compared to controls. Longitudinal follow-up will be important to determine whether the presence of inflammation predicts cognitive decline.

## INTRODUCTION

Evidence for inflammatory changes in Parkinson’s disease (PD) has been growing, with both innate and adaptive immune systems implicated. Post-mortem studies in brain tissue from subjects with PD reveal the presence of activated microglia and T-cells^1-4^ and evidence of immunoglobulin deposition^5^. Alterations in immune cell populations have been detected in living PD subjects, with most consistent findings pointing to alterations in T-cell and monocyte populations^6-11^. These changes may vary with disease duration or severity, although longitudinal studies to assess for changes in immune cells over time are lacking. Peptides derived from alpha-synuclein (αsyn), the key protein that aggregates in PD and the primary component of Lewy Bodies, can activate T-cells from PD patients^12^. Pro-inflammatory cytokines and chemokines are elevated in blood and cerebrospinal fluid (CSF) specimens from subjects with PD^13-26^. Brain imaging techniques using ligands against translocator protein 18kd (TSPO) as a marker for neuroinflammation have pointed to increased TSPO signal in PD patients^27-32^. Several animal PD models have demonstrated consistent inflammatory changes, and manipulation of inflammation can alter neurodegeneration in animal models^33-35^. Epidemiologic studies have suggested that immunomodulation may alter the course of PD: some data suggest that ibuprofen is protective against the development of PD^36-38^, while treatment of inflammatory bowel disease with anti-TNF biologics is associated with reduced PD risk^39 40^. Despite the wealth of information pointing to inflammatory activation in PD, many gaps remain in our understanding of the role of inflammation in the pathophysiology of PD.

An important question is whether inflammation contributes to the onset of PD pathogenesis or arises later in the course of the disease, in response to neurodegeneration. Genetic studies have linked polymorphisms in HLA genes to PD risk^41-44^, while several genes causal for PD, including *LRRK2, PRKN*, and *PINK1*, play a role in immune cells^34^, supporting an early role of the immune system. Association of certain infections with PD risk also point to a causal role for inflammation in PD pathogenesis^45^. Interestingly, some infections can stimulate αsyn levels^46 47^. T-cell changes are most prominent early and may even precede disease onset^48^. While these observations suggest a role of inflammation early in PD, most studies of inflammatory markers in human cohorts have examined a mix of subjects with a wide range of disease severity and duration and included patients already on treatment for PD. This is potentially problematic because dopaminergic drugs can alter immune responses^49^.

Another gap in understanding the role of inflammation is whether inflammation is associated with clinical features or predicts the subsequent course of disease, including cognitive decline. While many human studies have looked at inflammation in PD, these studies have been primarily performed in PD subjects with more advanced disease and lack longitudinal follow-up with clinical assessment and collection of inflammatory measures. Most studies also do not include detailed cognitive assessments. Additionally, there is a lack of studies that evaluate multiple inflammatory measures in the same cohort of patients.

To address these gaps in our understanding of the role of neuroinflammation in disease, we enrolled subjects with newly diagnosed, untreated PD (“*de novo*”) and age-matched healthy controls in a longitudinal study designed to comprehensively measure markers of inflammation and track clinical outcomes. Subjects enrolled in the study were enrolled within two years of PD diagnosis. Clinical evaluation of this cohort included PD-related scales and extensive cognitive testing consistent with the Level II diagnostic mild cognitive impairment (MCI) assessment criteria recommended by the Movement Disorders Society (MDS) task force^50^. Inflammatory measurements included plasma and CSF cytokine and chemokine measures, TSPO PET imaging, and peripheral immune cell phenotyping using flow cytometry. Here we describe clinical features and measures of inflammation in this cohort at the time of entry into the study.

## METHODS

### Standard protocol approvals, registrations, and patient consents

Participants were enrolled between November 2018 and December 2021 at the University of Alabama at Birmingham. The study was approved by the Institutional Review Board at UAB, and full written informed consent was obtained on each participant.

### Participants

Fifty-eight subjects with newly diagnosed PD and 62 age- and sex-matched healthy controls were enrolled in the Alabama Udall cohort. Inclusion criteria for PD subjects included the diagnosis of PD by United Kingdom Brain Bank criteria^51^ within two years of enrollment, age 40 or older at time of PD diagnosis, Hoehn and Yahr stage I through III, and no prior treatment with PD medications. Exclusion criteria for PD subjects included atypical Parkinsonian syndromes, clinical diagnosis of dementia, PD diagnosis greater than two years, previous treatment with PD medications, and the presence of other clinically significant neurological disorders. Inclusion criteria for control subjects were age 40 or greater, no current diagnosis of PD or other significant neurological disorder, no history of PD in first-degree blood relatives, and no more than 3 positive responses on the PD Screening Questionnaire^52^. Exclusion criteria for all participants included significant autoimmune or inflammatory disorders, active treatment with immunosuppressant therapy, and serious comorbidity that may interfere with study participation.

### Clinical Evaluation

After consent, participants underwent an extensive battery of clinical assessment over two visits. During the first visit, clinical assessment included demographics, general medical history, immune disorders and treatment questionnaire, vaccination history, prior and concomitant medication history, family history, behavioral history, PD Screening Questionnaire, vital signs, neurological examination, Epworth Sleepiness Scale, Hamilton Anxiety and Depression scales, Movement Disorder Society-United Parkinson’s Disease rating scale (MDS-UPDRS), Modified Schwab and England Activities of Daily Living, Montreal Cognitive Assessment (MoCA), Parkinson’s Disease Quality of Life Questionnaire (PDQ-39), Rapid Eye Movement Behavior Disorder Questionnaire, Pittsburgh Sleep Quality Index (PSQI), Scales for Outcomes in Parkinson’s Disease – Autonomic Dysfunction (SCOPA-AUT), and University of Pennsylvania Smell Identification Test (UPSIT). During the second visit, the participants underwent a comprehensive cognitive assessment that met the Level II diagnostic mild cognitive impairment (MCI) assessment criteria as recommended by the Movement Disorders Society (MDS) task force^50^. This test battery includes at least two tests across each of five cognitive domains: attention, language, memory, executive function, and visuospatial ability. To better describe patterns of cognitive performance, we also included separate domains for verbal memory and visual memory and included a processing speed domain. Tests performed were Wechsler Adult Intelligence Scale - Fourth Edition (WAIS-IV) Digit Span and Letter-Number Sequencing^53^; Hopkins Verbal Learning Test - Revised (HVLT- R)^54^; 10/36 Spatial Recall Test^55^; Judgment of Line Orientation (JLO)^56^; Hooper Visual Organization Test^57^; Boston Naming Test (BNT)^58^; Animal Naming^59^; Delis-Kaplan Executive Function System (DKEFS) Color/Word Interference^60^; and Trail Making Test (TMT)^61^. Normally distributed z-scores for each cognitive domain were calculated using the best available normative mean and standard deviation. Normative groups were stratified by age and education (when possible). At least two cognitive measures were used for each cognitive domain. Therefore, for each of the cognitive domains, an average z-score for the combined measures is used to represent an aggregated domain score. The cognitive composite score is the average of all domain z-scores.

### Biospecimen collection

At the baseline visit, blood samples were collected from each participant. Approximately 45 ml of blood was obtained and processed for flow cytometry studies, complete blood count, peripheral blood mononuclear cell preparation (PBMCs), plasma collection, DNA isolation, and TSPO SNP genotyping. Unused plasma and DNA samples were banked at the UAB Center for Clinical and Translational Science (CCTS) Sample Processing and Analysis Network (SPAN) and at the NINDS BioSEND program.

While not required for participation, some subjects also underwent CSF collection at baseline. CSF samples were banked at the CCTS SPAN until processed for cytokine/chemokine analysis. CSF was also banked at the NINDS BioSEND program.

### Cytokine/chemokine analysis

Plasma and CSF cytokine and chemokine levels were measured using the V-PLEX Proinflammatory Panel 1 and the V-PLEX Chemokine Panel 1 using the MSD SECTOR Imager 2400 through the UAB Diabetes Research Center Human Physiology Core.

### Flow cytometry

Human TruStain FcX (BioLegend, San Diego, CA) was used to block 500 μl of fresh peripheral blood prior to incubation with fluorochrome-conjugated monoclonal antibodies to monocyte, T-cell, and B-cell surface markers. RBC Red blood cells (RBC) were lysed using RBC Lysis Buffer (BioLegend San Diego, CA). After washing, cells were fixed with 2% paraformaldehyde. Cell viability was measured using the Aqua Live/Dead Kit (ThermoFisher Scientific). Antibodies used for surface markers were from BioLegend: anti-CD45 Pacific Blue (clone HI30); anti-CD3 Brilliant Violet 605/Brilliant Violet 650 (clone OKT3); anti-CD4 PE (clone OKT4); anti-CD4 eFluor 450 (clone OKT4); anti-CD8α FITC (clone HIT8a); anti-CD14 FITC (clone HCD14); anti-CD16 APC (clone 3G8); anti-CD25 PE/PerCP-Cyanine5.5 (clone M-A251); anti-CD19 Brilliant Violet 650 (clone HIB19); and anti-CD127 APC-Cy7 (clone A019D5). All antibodies were diluted to 1:100 and incubation time was 20-30 minutes at room temperature. Flow cytometry was performed on a FACSymphony (BD Biosciences). FlowJo software (Tree Star, Inc, Ashland, OR) was used for data analysis, as previously described ^62^.

### PET imaging

A 60-minute ^18^F-DPA-714 dynamic PET scan was acquired for each eligible subject in the baseline cohort. As the rs6971 single nucleotide polymorphism (SNP) is associated with TSPO radioligand binding affinity^63^, TSPO SNP genotyping was measured for every subject prior to PET, and those with low TSPO binding affinity were excluded from PET scans. Out of the 71 subjects who were PET-eligible, two subjects declined being scanned, one had a scan terminated early due to claustrophobia, and one subject who was scanned had image data unavailable due to scanner malfunction. A total of 67 subjects were successfully acquired for PET imaging. All subjects were scanned with a GE Signa PET/MR scanner. A sagittal T1-weighted scan was acquired shortly after the end of PET acquisition for brain parcellation. PET images were reconstructed with ordered subset expectation maximization with the zero-echo-time (ZTE) MRI-based attenuation correction^64^. All dynamic frames of the PET scan were co-registered to the last frame to reduce the patient motion during the acquisition. To extract the time-activity curves (TACs) for volumes of interest (VOIs), the brain parcellation was performed with FreeSurfer (v.7.1.1, Martinos Center, Boston MA)^65^ and then applied to the PET data. Partial volume correction was performed with the Geometric Transformation Matrix method in the PETPVC toolbox^66 67^. As FreeSurfer does not provide segmentation of the substantia nigra (SN), we have developed an in-house algorithm for fully automatic SN segmentation (see Supplemental Methods).

A total of ten brain regions were selected as the target VOI, including putamen, caudate nucleus, substantia nigra, hippocampus, thalamus, brainstem, frontal cortex, temporal cortex, parietal cortex, and occipital cortex. For each region, simplified reference tissue model (SRTM) was used to estimate the binding potential (BP), with the cerebellum cortex serving as the reference region ^68 69^. The binding potential was constrained to be non-negative for the SRTM optimization.

### Statistical Analysis

Characteristics of the study sample were described using means and SDs for continuous variables and frequencies (percentages) for categorical variables. Differences between groups were analyzed using independent samples *t* tests for continuous variables and Pearson chi-square tests (or Fisher’s exact tests) for categorical variables. For cytokine and chemokine comparisons, those samples with levels below the detectable range were considered “0” in the analysis. Similar findings were observed when these values were excluded or when they were replaced with detection limits.

Linear regression analysis was used to examine the associations of each regional ^18^F-DPA-714 binding potential with disease status, sex, and TSPO genotype. Correlations between each regional ^18^F-DPA-714 binding potential and other variables (clinical, cognitive, cytokine/chemokine, and immune cell phenotyping measures) were examined in the PD subjects only using linear regression analysis, adjusting for TSPO genotype. Results were considered statistically significant at p<0.05.

Statistical analyses were performed using version 4.2.0 Patched of the R programming environment (https://www.R-project.org).

### Data Sharing

Clinical assessment data have been uploaded to the NINDS Data Management Resource. Plasma, DNA, and CSF samples have been stored at the NINDS BioSEND repository. Additional data are available to qualified investigators on request.

## RESULTS

### Referral data

235 potential subjects were referred for this study by end of December 2022 (Supp. Fig. 1). 178 were eligible after screening. Out of the eligible candidates, 125 consented for the study, 4 were in queue for screening visit, and 49 refused to participate. 120 completed the enrollment visits, and 5 were screen failures at their enrollment visit.

### Baseline data

Among the 120 subjects enrolled, 58 subjects had PD, and 62 were controls. Baseline demographics, medical history, and family history were comparable between groups (Supp. Tables 1-3). Controls did differ from PD subjects for a subset of vaccinations, with PD having a higher rate of tetanus, diphtheria, and pertussis vaccination and a lower rate of COVID-19 vaccinations at time of enrollment (Supp. Table 4).

Among the clinical scale assessments, PD subjects showed significant differences in UPSIT, Schwab and England Activities of Daily Living, SCOPA-AUT, and MDS-UPDRS total scores (Table 1).

**Table 1.** Clinical assessment scores in patient cohort at baseline.

|  | **PD**<br>**N=58** | **Control**<br>**N=62** | ***p*** | **N** |
| --- | --- | --- | --- | --- |
| **Age** | 66.1 (8.55) | 64.1 (9.10) | 0.180 | 120 |
| **Gender** | 25 (43.1%) F | 34 (54.8%) F | 0.270 | 120 |
|  | 33 (56.9%) M | 28 (45.2%) M |  |  |
| **PDQ-39 Total** | 17.7 (16.5) | n/a | n/a | 57 |
| **MoCA total points** | 25.5 (3.73) | 27.5 (1.73) | <0.001 | 115 |
| **UPSIT Total Score** | 22.8 (7.84) | 32.4 (5.03) | <0.001 | 119 |
| **Schwab & England ADL Score** | 91.4 (10.2) | 97.9 (5.17) | <0.001 | 120 |
| **Epworth Sleepiness Scale Total** | 5.67 (4.21) | 5.10 (3.50) | 0.419 | 120 |
| **HAM-A Total Score** | 3.28 (3.09) | 2.61 (4.96) | 0.378 | 120 |
| **HAM-D Total Score** | 2.64 (2.63) | 3.16 (2.38) | 0.256 | 120 |
| **SCOPA-AUT Score** | 10.4 (6.63) | 5.85 (4.29) | <0.001 | 120 |
| **PSQI Total** | 5.14 (3.70) | 5.35 (3.43) | 0.740 | 120 |
| **UPDRS Total** | 43.4 (14.8) | 8.66 (5.89) | <0.001 | 119 |

Despite being within two years of PD diagnosis, PD subjects did show cognitive dysfunction, as determined by the cognitive battery. PD subjects showed worse performance on the MoCA (Table 1). PD subjects also showed significant differences in executive function, visuospatial ability, and cognitive composite at baseline compared to controls (Table 2).

**Table 2.** Cognitive data in patient cohort at baseline.

| **Cognitive Variable** | **PD**<br>**N=56** | **Control**<br>**N=60** | ***p*** | **N** |
| --- | --- | --- | --- | --- |
| **Attention** | .31 (.89) | .52 (.78) | 0.176 | 115 |
| **Verbal Memory** | 0.00 (1.07) | 0.25 (0.92) | 0.171 | 116 |
| **Visual Memory** | -0.01 (0.90) | 0.30 (0.82) | 0.054 | 115 |
| **Visuospatial** | 0.03 (0.93) | 0.34 (0.72) | 0.046 | 116 |
| **Language** | 0.34 (0.73) | 0.46 (0.66) | 0.348 | 115 |
| **Processing Speed** | 0.10 (0.79) | 0.26 (0.58) | 0.234 | 115 |
| **Executive Function** | 0.05 (0.91) | 0.36 (0.72) | 0.044 | 115 |
| **Cognitive Composite** | 0.14 (0.60) | 0.36 (0.47) | 0.033 | 114 |

Note. Values for Control and PD are mean z score (SD).

### Inflammatory measures in biospecimens

We measured cytokines and chemokines in the plasma of all control and PD subjects and in the CSF of a smaller subset of participants who agreed to undergo lumbar puncture (n= 19 controls and 25 PD subjects). We excluded those cytokines and chemokines in which levels for these molecules were below detection for >30% of subjects. Five molecules were excluded from the plasma analysis, and 9 were excluded from the CSF analysis. No significant differences were observed in chemokines and cytokines in the plasma specimens between control and PD participants, but an increase in plasma macrophage inflammatory protein-1β (MIP1β; also known as chemokine ligand 4 [CCL4]) among PD subjects compared to controls was noted (p = 0.054; Table 3). Examination of chemokines and cytokines in the CSF demonstrated statistically significant increases in macrophage inflammatory protein-1α (MIP1α/CCL3; p = 0.043) and thymus- and activation-regulated chemokine (TARC/CCL17) in PD subjects and an increased trend in macrophage-derived chemokine (MDC/CCL22; p = 0.025) in PD subjects compared to controls (p=0.073; Table 3).

**Table 3.** Baseline cytokine levels in plasma and CSF.

|  | **Control**<br>**N=62** | **PD**<br>**N=56** | ***p*** | **N** |
| --- | --- | --- | --- | --- |
| ***plasma*** |  |  |  |  |
| Eotaxin | 192 (74.1) | 207 (79.7) | 0.302 | 118 |
| Eotaxin-3 | 17.9 (13.7) | 18.8 (9.85) | 0.678 | 117 |
| IP-10 | 265 (128) | 327 (256) | 0.109 | 118 |
| MCP-1 | 87.9 (36.0) | 146 (474) | 0.368 | 118 |
| MCP-4 | 123 (75.9) | 127 (57.4) | 0.734 | 118 |
| MDC | 1040 (378) | 1191 (1032) | 0.306 | 118 |
| MIP-1α | 10.4 (3.55) | 55.1 (231) | 0.153 | 118 |
| MIP-1β | 68.8 (23.3) | 78.4 (29.8) | 0.054 | 118 |
| TARC | 268 (224) | 265 (152) | 0.912 | 118 |
| IL-10 | 0.28 (0.14) | 0.37 (0.42) | 0.118 | 118 |
| IL-8 | 7.26 (4.71) | 6.47 (2.19) | 0.243 | 118 |
| TNF-α | 1.40 (0.41) | 1.49 (0.51) | 0.304 | 118 |
| IFN-γ | 5.62 (3.54) | 5.25 (3.25) | 0.555 | 118* |
| IL-6 | 1.08 (0.79) | 1.07 (0.88) | 0.916 | 118* |
| ***CSF*** |  |  |  |  |
|  | **Control**<br>**N=19** | **PD**<br>**N=25** | ***p*** | **N** |
| IL-2 | 0.23 (0.13) | 0.87 (2.17) | 0.156 | 44 |
| IL-8 | 41.9 (10.3) | 40.2 (12.0) | 0.619 | 44 |
| Eotaxin | 16.5 (10.1) | 19.6 (6.73) | 0.254 | 44* |
| IP-10 | 341 (195) | 419 (265) | 0.270 | 44* |
| MCP-1 | 342 (66.7) | 324 (107) | 0.478 | 44* |
| MDC | 9.68 (6.55) | 13.2 (5.73) | 0.073 | 44* |
| MIP-1α | 5.43 (2.58) | 6.90 (1.81) | 0.043 | 44* |
| MIP-1β | 9.25 (2.17) | 10.7 (4.07) | 0.139 | 44* |
| TARC | 2.48 (1.48) | 3.51 (1.42) | 0.025 | 44* |
| IL-6 | 0.93 (0.52) | 1.17 (0.71) | 0.195 | 44* |

\* A few subject samples had undetectable values for the measured cytokine or chemokine. These undetectable values were replaced with zeros.

In addition to cytokine and chemokine analyses, flow cytometry was performed in biospecimens obtained from participants. PD participants demonstrated an increase in CD4^+^ T non-regulatory cell percentages (CD4^+^CD25^+^CD127^+^; p = 0.048) with a decrease in CD4^+^ T regulatory (Tregs) cell percentages (CD4^+^CD25^+^CD127^+^; p = 0.038) compared to controls (Fig. 1a, b). Additionally, PD subjects showed a higher neutrophil to lymphocyte ratio (NLR) compared to controls (Fig. 1c; p = 0.004). No changes in monocyte, CD8^+^ T-cell, or B-cell percentages were observed at baseline (data not shown).

**Figure 1.**
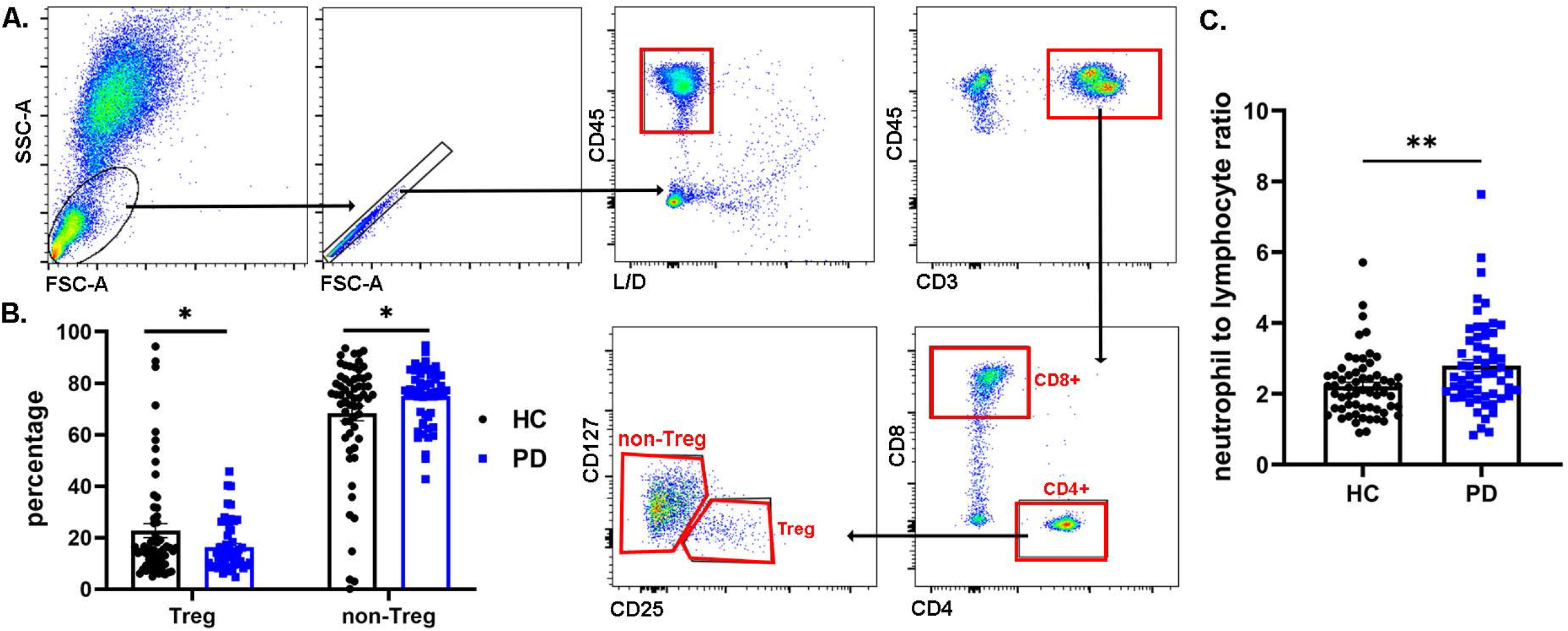
Alterations in T regulatory subset, T non-regulatory subset, and Neutrophil to lymphocyte ratio (NLR) in PD. **A**.Gating strategies for CD4^+^CD25^+^CD127-Treg and CD4^+^CD25^+^CD127^+^ non-Treg subsets. **B**.Percentages of Treg and non-Treg subsets from control (HC) and PD subjects shown as mean ± SEM. *p<0.05 (Student’s t-test with Welch’s correction). **C**.NLR shown as mean ± SEM. **p<0.01.

### PET imaging

Control and PD participants underwent TSPO SNP genotyping to determine if they were eligible for ^18^F-DPA-714 imaging. Among the 88 participants willing to undergo ^18^F-DPA-714 imaging, 14.8% were low affinity binders (LAB), 30.7% were mixed affinity binders (MAB), and 54.5% were high affinity binders (HAB). Only MAB and HAB subjects were imaged, with a total of 37 controls and 30 PD subjects who had ^18^F-DPA-714 imaging successfully completed. We observed a significant difference in unadjusted TSPO binding potential between control and PD participants at baseline in several brain regions (Fig. 2). Using linear regression to adjust for TSPO genotype and sex, we found that PD subjects had higher ^18^F-DPA-714 binding potential in the putamen, thalamus, substantia nigra, temporal cortex, parietal cortex, and occipital cortex (Supp. Table 5; p=0.013 for putamen, p=0.022 for thalamus, p=0.045 for SN, p=0.027 for temporal cortex, p=0.012 for parietal cortex, and p=0.00093 for occipital cortex). No significant differences in ^18^F-DPA 714 binding potential were noted in the hippocampus, brainstem, caudate, or frontal cortex after adjustment for TSPO genotype and sex. As expected, TSPO genotype affected ^18^F-DPA-714 binding potential independent of diagnosis or sex in several brain regions (Supp. Table 5; p=0.0085 for putamen, p=0.0073 for brainstem, p=0.00043 for frontal cortex, p=0.023 for parietal cortex). Interestingly, we also observed a sex difference between male and female in ^18^F-DPA 714 binding potential independent of diagnosis and TSPO genotype in the putamen and thalamus (Supp. Table 5; p=0.011 in putamen, p=0.0043 in thalamus).

**Figure 2.**
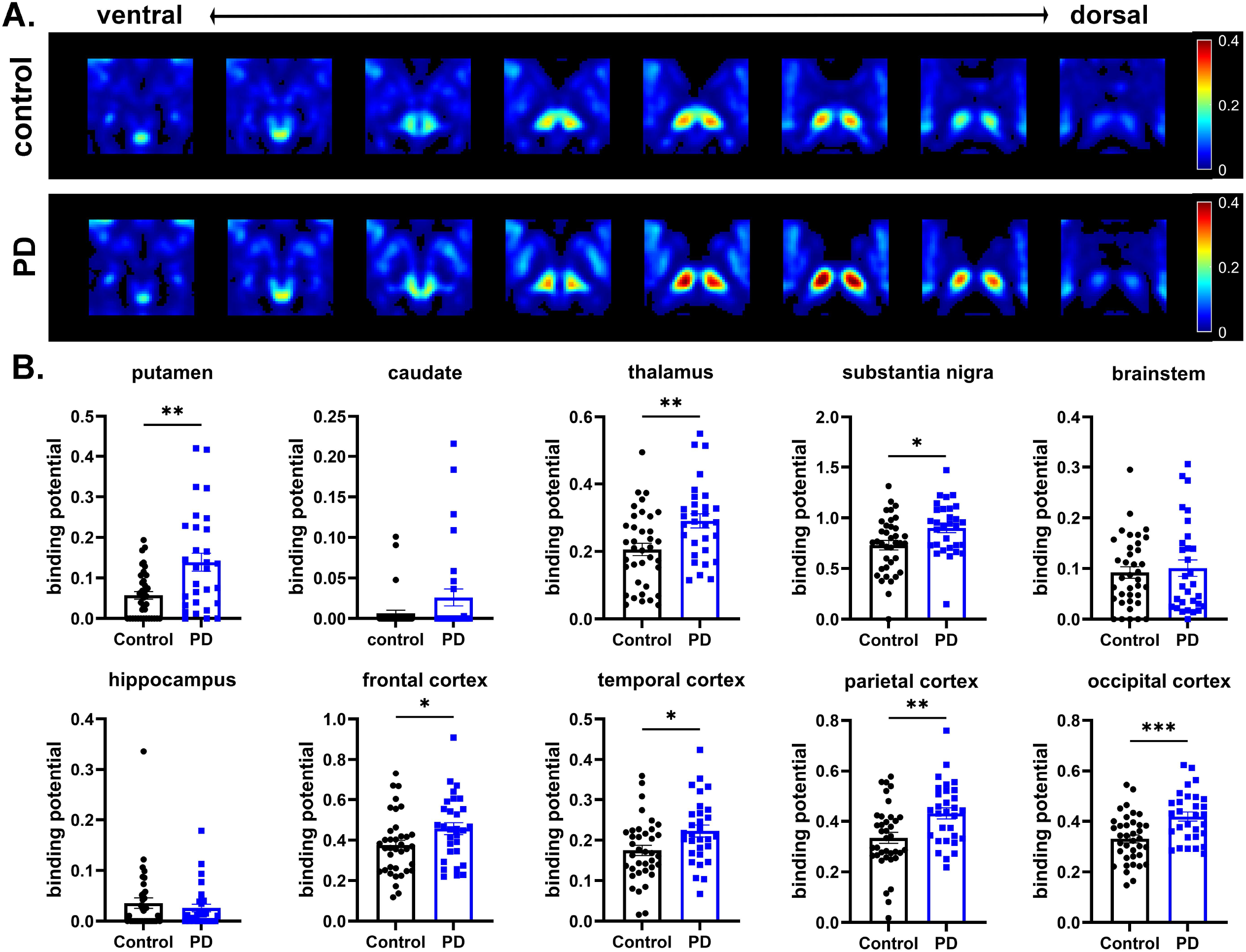
PD subjects show higher ^18^F-DPA-714 binding potential in the putamen, thalamus, substantia nigra, and cortical brain regions. **A**.Comparison of the parametric images of binding potential in the striatal and thalamus area between the control and PD cohorts. Mean of the BP images for HAB subjects is shown here. **B**.Raw values for binding potential in control and PD subjects that is not adjusted for sex or TSPO genotype. *p<0.05, **p<0.01, ***p<0.001 (Student *t*-test with Welch’s correction). Using linear regression to adjust for TSPO genotype and sex, TSPO binding potential is higher in PD subjects in the putamen, thalamus, substantia nigra, and several cortical brain regions.

Given the significant differences in central inflammation as noted by ^18^F-DPA-714 imaging, we next performed correlation analyses to determine whether specific clinical measures, plasma or CSF cytokines and chemokines, or immune cell changes were associated with the ^18^F-DPA-714 changes among the PD subjects. Using univariate analysis adjusting for TSPO genotyping, we observed significant correlations between ^18^F-DPA-714 binding potential in the thalamus with several cognitive domains, including visuospatial, language, verbal memory, and composite cognitive domain scores (Supp. Table 6). In terms of cognitive domains that showed correlations with ^18^F-DPA-714 binding potential in multiple brain regions, we noted significant correlations between the visuospatial domain and ^18^F-DPA-714 binding potential in the caudate, thalamus, and brainstem and between language domain scores and ^18^F-DPA-714 binding potential in the putamen and thalamus. As expected with the large number of variables examined in this discovery study, associations were no longer significant after full adjustment for multiple comparisons.

^18^F-DPA-714 binding potential also significantly correlated with certain inflammatory measures across several brain regions, including a subset of plasma and CSF cytokines and chemokines among PD participants. Specifically, we observed significant associations between plasma eotaxin 3/CCL26 and ^18^F-DPA-714 binding potential for several brain regions including the putamen, thalamus, and hippocampus (Supp. Table 6). We also noted significant correlations between CSF macrophage-derived chemokine (MDC/CCL22) and ^18^F-DPA-714 binding potential in the putamen, thalamus, hippocampus, and brainstem (Supp. Table 6). Once again, these associations were no longer significant after full adjustment for multiple comparisons. No significant correlations were observed between PET measures and immune cell phenotyping by flow cytometry, except for NLR with ^18^F-DPA-714 binding potential in the substantia nigra (Supp. Table 6).

## DISCUSSION

In this study, we enrolled subjects with newly diagnosed PD and age-matched controls in a longitudinal study designed to comprehensively measure markers of inflammation and track clinical outcomes. Baseline ^18^F-DPA-714 imaging pointed to evidence for central inflammation in newly diagnosed PD patients within two years of diagnosis and not treated with PD medications. PD subjects at baseline also demonstrated alterations in executive function, visual memory, and cognitive composite domains compared to controls. PET imaging changes, particularly in the thalamus, correlated with certain cognitive domain subscores and composite scores and correlated with a subset of plasma and CSF chemokines.

Our study demonstrates that TSPO binding is elevated in early, untreated PD. Several prior studies have examined TSPO binding in PD populations^27-32^, but all these enrolled smaller numbers of PD subjects from a broad range of PD stage and duration and many who were already on active PD treatment. Our data demonstrate clearly that increased TSPO binding is present in PD independent of any treatment effects. In addition, our multimodal study provides further evidence that TSPO signal as measured by ^18^F-DPA-714 is a marker of central inflammation, as we found correlations of ^18^F-DPA-714 signal with chemokine measures. In particular, ^18^F-DPA-714 signal was associated with MDC/CCL22. This attractant for monocytes, Th2 T-cells, and T regulatory cells has been shown to be produced by microglia in experimental autoimmune encephalitis models^70^. The correlation between ^18^F-DPA-714 and MDC/CCL2 suggests that the ^18^F-DPA-714 signal is partly secondary to microglial activation, although the ^18^F-DPA-714 signal could also mark peripheral immune cell infiltration in response to MDC/CCL2 release. Indeed, the finding that increased ^18^F-DPA-714 binding was observed in most brain regions examined in PD subjects fits with this concept of peripheral immune cell infiltration early in PD^71^.

With our baseline dataset, we performed a discovery-based univariate correlation analyses of the PET data with clinical measures to find whether any associations may be present at baseline within the PD group. The most interesting associations were found within the cognitive domains. In particular, ^18^F-DPA-714 binding potential was associated with the visuospatial domain subscore and the cognitive composite score in several brain regions. Another interesting observation was that the ^18^F-DPA-714 binding potential within the thalamus correlated with several cognitive domains, including verbal memory, visuospatial, and language domains. This observation suggests an important role for inflammation in the thalamus regarding cognitive findings early in PD.

We also found an intriguing observation that ^18^F-DPA-714 binding potential differs between males and females, with females showing reduced binding potential compared to males, independent of disease. This finding suggests that central inflammation as detected by TSPO PET differs between males and females. We have previously noted sex-related differences in the inflammatory response in PD, including sex differences in monocyte activation in PD^7^. Such sex-based differences in the immune response could point to potential reasons for the differences in PD prevalence observed between male and females. Of note, Tuisku *et al*. evaluated the effect of age and sex on ^11^C-PBR28 TSPO imaging and observed that healthy females had higher total volume of distribution (Vt) compared to healthy males, but that TSPO Vt increased with age in males^72^. In our study, both males and females were mostly older than those studied by Tuisku *et al*., which, in addition to structural differences between ^11^C-PBR28 and ^18^F-DPA-714, may account for the differences in the measured effects of sex on TSPO imaging.

A limitation of the use of TSPO PET imaging techniques to measure brain inflammation is that it is unclear what cell type is responsible for the TSPO binding signal. For many years, TSPO was thought to be specific to microglial activation, yet TSPO can also be bound by other inflammatory cells, including astrocytes and peripheral immune cells^73-76^. More recent studies in rodent models have suggested that TSPO signal may be related more to microglial number than microglial activation^77^. While our observations suggest that increased TSPO signal reflects central inflammation and correlates with cognitive measures, the TSPO signal we observed is found in multiple brain regions and does not offer clear insight into the cell types that may be responsible.

Additionally, second generation TSPO ligands, including ^18^F-DPA-714, are affected by the SNP rs6971 genotype^63^. We have previously shown that imaging with ^18^F-DPA-714 PET can reveal the difference between TSPO binding in HAB and MAB normal controls^78^. When we examine our PET data by genotype, it is apparent that much of the difference between control and PD participants is driven by observations in the HAB subjects (Supp. Table 7). The MAB group was smaller (only about 30% of the overall sample) and the mean TSPO binding signal among MAB was not much higher than background levels. When analyzed separately, the MAB group showed no statistical difference between control and PD subjects (Supp. Table 7). From a practical point of view, this suggests that TSPO PET is likely to be most useful in HAB subjects. As HAB subjects are only half the population in our sample, alternative ligands not affected by this TSPO SNP are needed to bring PET imaging into the clinical arena.

Previous studies of chemokine and cytokine levels in plasma or CSF have pointed to elevations in several pro-inflammatory molecules, although studies vary in the degree and particular cytokines or chemokines observed^13-26^. In the blood, the most consistent changes have been noted in IL-1β and some studies have also pointed to changes in IL-2, IFN-γ, and TNF-α^26^. Two recent studies have evaluated serum cytokines in early-stage PD subjects^13 25^. In the ICICLE study, PD subjects at early stages showed increased TNF-α, IL-1β, IL-2, and IL-10 in sera compared to controls, with subjects showing a “pro-inflammatory” profile having faster progression on the mini mental status exam (MMSE) over 36 months^25^. In another study measuring these cytokines in early PD, Ahmad Rastegar *et al* found that MIP-1α and MCP-1 predicted motor decline at two years in a cohort of PD patients with and without LRRK2 mutations^13^. We found no significant differences in plasma cytokine levels between controls and PD subjects, although we did see a near statistically significant increase in MIP-1β and a trend in MIP-1α. Potential differences between our findings and those studies performed at earlier stages include the smaller sample size in our group and the inclusion of a substantial number of LRRK2 mutation carriers in the Ahmad Rastegar study.

In the CSF, we observed significant increases in MIP-1α and TARC in PD subjects compared to controls. Previous studies have shown increases among different cytokines and chemokines, and while specific inflammatory markers have differed between studies, most consistent changes have been noted in IL-1β and IL-6 in the CSF of PD patients^14 16 19 21 26^. In our cohort, we did see a non-significant increase in IL-6, and IL-1β levels were below detection in a significant subset of subjects. Potential reasons for differences in our cohort and other published cohorts include differences in sample size, differences in disease duration and severity, and use of PD medication in other cohorts. Additionally, diurnal variations in CSF inflammatory molecules are greater in PD subjects compared to controls^79^.

In the Udall cohort, we observed an increase in the NLR in PD subjects; this marker of systemic inflammation has been observed in other PD cohorts^80^. We also saw a significant decrease in CD4^+^ Tregs in PD subjects compared to controls. CD4^+^ Tregs are defined by the surface markers CD25^+^CD127^-^ and have potent anti-inflammatory and immunosuppressive properties^81-83^. In contrast, CD4^+^ non-Tregs, defined by surface markers CD25^+^CD127^+^, were significantly increased in PD subjects compared to controls. CD4^+^ non-Tregs have pro-inflammatory properties^82^. This decrease in Tregs and increase in non-Tregs suggests a deficient anti-inflammatory response, which may promote the chronic inflammation associated with PD. These findings support previous observations of decreased percentages of Tregs in PD participants^6 10 11^.

In conclusion, we found elevated ^18^F-DPA-714 binding in newly diagnosed PD patients compared to control subjects, and this signal correlated with cognitive testing and a subset of plasma and CSF cytokines. This study supports the conclusion that central inflammation is observed early in the disease process in PD, is independent of treatment for PD, and is correlated with cognitive features and peripheral markers of inflammation. Future directions include longitudinal follow-up of this cohort to determine if certain inflammatory changes are predictive of clinical progression, in particular regarding cognitive function. We will also continue to collect biospecimens annually to determine if the inflammatory measures change over time in PD subjects.

## Acknowledgements

This work was supported by the National Institutes of Health P50NS108675.

## Author Roles

TAY, AG, ENB, JM, and DGS designed research; TAY, AWA, NS, LR, CC, HH, HQ executed research; TAY, Y-H F, AG, RK, YZ, HQ analyzed data; TAY, Y-H F, AG, RK, YZ, ENB, DGS wrote the manuscript; TAY, Y-H F, AG, NS, RK, YZ, HQ, ENB, DGS edited the final manuscript.

## References

1. Brochard V, Combadiere B, Prigent A, et al. Infiltration of CD4+ lymphocytes into the brain contributes to neurodegeneration in a mouse model of Parkinson disease. J Clin Invest 2009;119(1):182-92. doi: 10.1172/JCI36470 [published Online First: 2008/12/24]

2. McGeer PL, Itagaki S, Akiyama H, et al. Rate of cell death in parkinsonism indicates active neuropathological process. Ann Neurol 1988;24(4):574–6. doi: 10.1002/ana.410240415 [published Online First: 1988/10/01]

3. McGeer PL, Itagaki S, Boyes BE, et al. Reactive microglia are positive for HLA-DR in the substantia nigra of Parkinson’s and Alzheimer’s disease brains. Neurology 1988;38(8):1285–91. doi: 10.1212/wnl.38.8.1285 [published Online First: 1988/08/01]

4. Sommer A, Marxreiter F, Krach F, et al. Th17 Lymphocytes Induce Neuronal Cell Death in a Human iPSC-Based Model of Parkinson’s Disease. Cell Stem Cell 2018;23(1):123–31 e6. doi: 10.1016/j.stem.2018.06.015 [published Online First: 2018/07/07]

5. Orr CF, Rowe DB, Mizuno Y, et al. A possible role for humoral immunity in the pathogenesis of Parkinson’s disease. Brain 2005;128(Pt 11):2665-74. doi: 10.1093/brain/awh625 [published Online First: 2005/10/13]

6. Alvarez-Luquin DD, Arce-Sillas A, Leyva-Hernandez J, et al. Regulatory impairment in untreated Parkinson’s disease is not restricted to Tregs: other regulatory populations are also involved. J Neuroinflammation 2019;16(1):212. doi: 10.1186/s12974-019-1606-1 [published Online First: 2019/11/13]

7. Carlisle SM, Qin H, Hendrickson RC, et al. Sex-based differences in the activation of peripheral blood monocytes in early Parkinson disease. NPJ Parkinsons Dis 2021;7(1):36. doi: 10.1038/s41531-021-00180-z [published Online First: 2021/04/15]

8. Grozdanov V, Bliederhaeuser C, Ruf WP, et al. Inflammatory dysregulation of blood monocytes in Parkinson’s disease patients. Acta Neuropathol 2014;128(5):651–63. doi: 10.1007/s00401-014-1345-4 [published Online First: 2014/10/07]

9. Jiang S, Gao H, Luo Q, et al. The correlation of lymphocyte subsets, natural killer cell, and Parkinson’s disease: a meta-analysis. Neurol Sci 2017;38(8):1373–80. doi: 10.1007/s10072-017-2988-4 [published Online First: 2017/05/13]

10. Saunders JA, Estes KA, Kosloski LM, et al. CD4+ regulatory and effector/memory T cell subsets profile motor dysfunction in Parkinson’s disease. J Neuroimmune Pharmacol 2012;7(4):927–38. doi: 10.1007/s11481-012-9402-z [published Online First: 2012/10/12]

11. Kustrimovic N, Rasini E, Legnaro M, et al. Dopaminergic receptors on CD4+ T naive and memory lymphocytes correlate with motor impairment in patients with Parkinson’s Disease. Sci Rep 2016;6:33738. doi: 10.1038/srep33738

12. Sulzer D, Alcalay RN, Garretti F, et al. T cells from patients with Parkinson’s disease recognize alpha-synuclein peptides. Nature 2017;546(7660):656–61. doi: 10.1038/nature22815 [published Online First: 2017/06/22]

13. Ahmadi Rastegar D, Ho N, Halliday GM, et al. Parkinson’s progression prediction using machine learning and serum cytokines. NPJ Parkinsons Dis 2019;5:14. doi: 10.1038/s41531-019-0086-4 [published Online First: 2019/08/03]

14. Blum-Degen D, Muller T, Kuhn W, et al. Interleukin-1 beta and interleukin-6 are elevated in the cerebrospinal fluid of Alzheimer’s and de novo Parkinson’s disease patients. Neurosci Lett 1995;202(1-2):17–20. doi: 10.1016/0304-3940(95)12192-7 [published Online First: 1995/12/29]

15. Brodacki B, Staszewski J, Toczylowska B, et al. Serum interleukin (IL-2, IL-10, IL-6, IL-4), TNFalpha, and INFgamma concentrations are elevated in patients with atypical and idiopathic parkinsonism. Neurosci Lett 2008;441(2):158–62. doi: 10.1016/j.neulet.2008.06.040 [published Online First: 2008/06/28]

16. Chen X, Hu Y, Cao Z, et al. Cerebrospinal Fluid Inflammatory Cytokine Aberrations in Alzheimer’s Disease, Parkinson’s Disease and Amyotrophic Lateral Sclerosis: A Systematic Review and Meta-Analysis. Front Immunol 2018;9:2122. doi: 10.3389/fimmu.2018.02122 [published Online First: 2018/10/05]

17. Hall S, Janelidze S, Surova Y, et al. Cerebrospinal fluid concentrations of inflammatory markers in Parkinson’s disease and atypical parkinsonian disorders. Sci Rep 2018;8(1):13276. doi: 10.1038/s41598-018-31517-z [published Online First: 2018/09/07]

18. Karpenko MN, Vasilishina AA, Gromova EA, et al. Interleukin-1beta, interleukin-1 receptor antagonist, interleukin-6, interleukin-10, and tumor necrosis factor-alpha levels in CSF and serum in relation to the clinical diversity of Parkinson’s disease. Cell Immunol 2018;327:77–82. doi: 10.1016/j.cellimm.2018.02.011 [published Online First: 2018/02/27]

19. Mogi M, Harada M, Narabayashi H, et al. Interleukin (IL)-1 beta, IL-2, IL-4, IL-6 and transforming growth factor-alpha levels are elevated in ventricular cerebrospinal fluid in juvenile parkinsonism and Parkinson’s disease. Neurosci Lett 1996;211(1):13–6. doi: 10.1016/0304-3940(96)12706-3 [published Online First: 1996/06/14]

20. Mogi M, Harada M, Riederer P, et al. Tumor necrosis factor-alpha (TNF-alpha) increases both in the brain and in the cerebrospinal fluid from parkinsonian patients. Neurosci Lett 1994;165(1-2):208–10. doi: 10.1016/0304-3940(94)90746-3 [published Online First: 1994/01/03]

21. Muller T, Blum-Degen D, Przuntek H, et al. Interleukin-6 levels in cerebrospinal fluid inversely correlate to severity of Parkinson’s disease. Acta Neurol Scand 1998;98(2):142–4. doi: 10.1111/j.1600-0404.1998.tb01736.x [published Online First: 1998/09/02]

22. Qin XY, Zhang SP, Cao C, et al. Aberrations in Peripheral Inflammatory Cytokine Levels in Parkinson Disease: A Systematic Review and Meta-analysis. JAMA Neurol 2016;73(11):1316–24. doi: 10.1001/jamaneurol.2016.2742 [published Online First: 2016/09/27]

23. Reale M, Iarlori C, Thomas A, et al. Peripheral cytokines profile in Parkinson’s disease. Brain Behav Immun 2009;23(1):55–63. doi: 10.1016/j.bbi.2008.07.003 [published Online First: 2008/08/06]

24. Scalzo P, Kummer A, Cardoso F, et al. Serum levels of interleukin-6 are elevated in patients with Parkinson’s disease and correlate with physical performance. Neurosci Lett 2010;468(1):56–8. doi: 10.1016/j.neulet.2009.10.062 [published Online First: 2009/10/28]

25. Williams-Gray CH, Wijeyekoon R, Yarnall AJ, et al. Serum immune markers and disease progression in an incident Parkinson’s disease cohort (ICICLE-PD). Mov Disord 2016;31(7):995–1003. doi: 10.1002/mds.26563

26. Zimmermann M, Brockmann K. Blood and Cerebrospinal Fluid Biomarkers of Inflammation in Parkinson’s Disease. J Parkinsons Dis 2022 doi: 10.3233/JPD-223277 [published Online First: 2022/06/07]

27. Iannaccone S, Cerami C, Alessio M, et al. In vivo microglia activation in very early dementia with Lewy bodies, comparison with Parkinson’s disease. Parkinsonism Relat Disord 2013;19(1):47–52. doi: 10.1016/j.parkreldis.2012.07.002 [published Online First: 2012/07/31]

28. Lavisse S, Goutal S, Wimberley C, et al. Increased microglial activation in patients with Parkinson disease using [(18)F]-DPA714 TSPO PET imaging. Parkinsonism Relat Disord 2021;82:29–36. doi: 10.1016/j.parkreldis.2020.11.011 [published Online First: 2020/11/27]

29. Ouchi Y, Yoshikawa E, Sekine Y, et al. Microglial activation and dopamine terminal loss in early Parkinson’s disease. Ann Neurol 2005;57(2):168–75. doi: 10.1002/ana.20338 [published Online First: 2005/01/26]

30. Terada T, Yokokura M, Yoshikawa E, et al. Extrastriatal spreading of microglial activation in Parkinson’s disease: a positron emission tomography study. Ann Nucl Med 2016;30(8):579–87. doi: 10.1007/s12149-016-1099-2 [published Online First: 2016/06/15]

31. Zhang PF, Gao F. Neuroinflammation in Parkinson’s disease: a meta-analysis of PET imaging studies. J Neurol 2022;269(5):2304–14. doi: 10.1007/s00415-021-10877-z [published Online First: 2021/11/02]

32. Liu S-Y, Qiao H-W, Song T-B, et al. Brain microglia activation and peripheral adaptive immunity in Parkinson’s disease: a multimodal PET study. Journal of Neuroinflammation 2022;19(1):209. doi: 10.1186/s12974-022-02574-z

33. Qin H, Buckley JA, Li X, et al. Inhibition of the JAK/STAT pathway protects against α-synuclein-induced neuroinflammation and dopaminergic neurodegeneration. J Neurosci 2016;36(18):5144–59. doi: 10.1523/jneurosci.4658-15.2016

34. Tansey MG, Wallings RL, Houser MC, et al. Inflammation and immune dysfunction in Parkinson disease. Nat Rev Immunol 2022 doi: 10.1038/s41577-022-00684-6 [published Online First: 2022/03/06]

35. Weiss F, Labrador-Garrido A, Dzamko N, et al. Immune responses in the Parkinson’s disease brain. Neurobiol Dis 2022;168:105700. doi: 10.1016/j.nbd.2022.105700 [published Online First: 2022/03/23]

36. Chen H, Jacobs E, Schwarzschild MA, et al. Nonsteroidal antiinflammatory drug use and the risk for Parkinson’s disease. Ann Neurol 2005;58(6):963–7. doi: 10.1002/ana.20682 [published Online First: 2005/10/22]

37. Chen H, Zhang SM, Hernan MA, et al. Nonsteroidal anti-inflammatory drugs and the risk of Parkinson disease. Arch Neurol 2003;60(8):1059–64. doi: 10.1001/archneur.60.8.1059 [published Online First: 2003/08/20]

38. Samii A, Etminan M, Wiens MO, et al. NSAID use and the risk of Parkinson’s disease: systematic review and meta-analysis of observational studies. Drugs Aging 2009;26(9):769–79. doi: 10.2165/11316780-000000000-00000 [published Online First: 2009/09/05]

39. Park S, Kim J, Chun J, et al. Patients with Inflammatory Bowel Disease Are at an Increased Risk of Parkinson’s Disease: A South Korean Nationwide Population-Based Study. J Clin Med 2019;8(8) doi: 10.3390/jcm8081191 [published Online First: 2019/08/11]

40. Peter I, Dubinsky M, Bressman S, et al. Anti-Tumor Necrosis Factor Therapy and Incidence of Parkinson Disease Among Patients With Inflammatory Bowel Disease. JAMA Neurol 2018;75(8):939–46. doi: 10.1001/jamaneurol.2018.0605 [published Online First: 2018/05/02]

41. Hamza TH, Zabetian CP, Tenesa A, et al. Common genetic variation in the HLA region is associated with late-onset sporadic Parkinson’s disease. Nat Genet 2010;42(9):781–5. doi: 10.1038/ng.642 [published Online First: 2010/08/17]

42. Kannarkat GT, Cook DA, Lee JK, et al. Common Genetic Variant Association with Altered HLA Expression, Synergy with Pyrethroid Exposure, and Risk for Parkinson’s Disease: An Observational and Case-Control Study. NPJ Parkinsons Dis 2015;1 doi: 10.1038/npjparkd.2015.2 [published Online First: 2015/01/01]

43. Nalls MA, Pankratz N, Lill CM, et al. Large-scale meta-analysis of genome-wide association data identifies six new risk loci for Parkinson’s disease. Nat Genet 2014;46(9):989–93. doi: 10.1038/ng.3043 [published Online First: 2014/07/30]

44. Wissemann WT, Hill-Burns EM, Zabetian CP, et al. Association of Parkinson disease with structural and regulatory variants in the HLA region. Am J Hum Genet 2013;93(5):984–93. doi: 10.1016/j.ajhg.2013.10.009 [published Online First: 2013/11/05]

45. Smeyne RJ, Noyce AJ, Byrne M, et al. Infection and Risk of Parkinson’s Disease. J Parkinsons Dis 2021;11(1):31–43. doi: 10.3233/JPD-202279 [published Online First: 2020/12/29]

46. Beatman EL, Massey A, Shives KD, et al. Alpha-Synuclein Expression Restricts RNA Viral Infections in the Brain. J Virol 2015;90(6):2767–82. doi: 10.1128/JVI.02949-15 [published Online First: 2016/01/01]

47. Stolzenberg E, Berry D, Yang, et al. A Role for Neuronal Alpha-Synuclein in Gastrointestinal Immunity. J Innate Immun 2017;9(5):456–63. doi: 10.1159/000477990 [published Online First: 2017/06/27]

48. Lindestam Arlehamn CS, Dhanwani R, Pham J, et al. α-Synuclein-specific T cell reactivity is associated with preclinical and early Parkinson’s disease. Nature communications 2020;11(1):1875. doi: 10.1038/s41467-020-15626-w [published Online First: 2020/04/22]

49. Li M, Zhou L, Sun X, et al. Dopamine, a co-regulatory component, bridges the central nervous system and the immune system. Biomedicine & pharmacotherapy = Biomedecine & pharmacotherapie 2022;145:112458. doi: 10.1016/j.biopha.2021.112458 [published Online First: 2021/12/01]

50. Litvan I, Goldman JG, Troster AI, et al. Diagnostic criteria for mild cognitive impairment in Parkinson’s disease: Movement Disorder Society Task Force guidelines. Mov Disord 2012;27(3):349–56. doi: 10.1002/mds.24893

51. Hughes AJ, Daniel SE, Kilford L, et al. Accuracy of clinical diagnosis of idiopathic Parkinson’s disease: a clinico-pathological study of 100 cases. J Neurol Neurosurg Psychiatry 1992;55(3):181–4.

52. Rocca WA, Maraganore DM, McDonnell SK, et al. Validation of a telephone questionnaire for Parkinson’s disease. J Clin Epidemiol 1998;51(6):517–23. doi: 10.1016/s0895-4356(98)00017-1 [published Online First: 1998/06/23]

53. Wechsler D, Psychological C, PsychCorp. WAIS-IV technical and interpretive manual. San Antonio, Tex.: Pearson 2008.

54. Brandt J, Benedict R. Hopkins Verbal Learning Test-Revised: Professional Manual. Florida: PAR 2001.

55. Rao SM, Cognitive Function Study Group N. A manual for the Brief Repeatable Battery of Neuropsychological Tests in Multiple Sclerosis. New York: National Multiple Sclerosis Society 1990.

56. Benton AI, Sivan AB, Hamsher K, et al. Contributions to Neuropsychological Assessment: A Clinical Manual - 2nd ed. New York, NY: Oxford 1994.

57. Hooper HE. Hooper Visual Organization Test. Los Angeles, CA: Western Psychological Services 1983.

58. Kaplan E, Goodlass H, Weintraub S. The Boston Naming Test. Philadelphia, PA: Lea & Febiger 1983.

59. Ruff RM, Light RH, Parker SB, et al. Benton Controlled Oral Word Association Test: reliability and updated norms. Archives Of Clinical Neuropsychology: The Official Journal Of The National Academy Of Neuropsychologists 1996;11(4):329–38.

60. Delis DC, Kaplan E, Kramer JH. Delis-Kaplin Executive Function System (D-KEFS): Examiner’s manual. San Antonio, TX: The Psychological Corporation 2001.

61. Spreen O, Strauss E. A compendium of neuropsychological tests: Administration, norms and commentary. 2nd edition. New York: Oxford University Press 1998.

62. Yan Z, Yang W, Parkitny L, et al. Deficiency of Socs3 leads to brain-targeted EAE via enhanced neutrophil activation and ROS production. JCI Insight 2019;5(9):e126520. doi: 10.1172/jci.insight.126520 [published Online First: 2019/04/03]

63. Owen DR, Yeo AJ, Gunn RN, et al. An 18-kDa translocator protein (TSPO) polymorphism explains differences in binding affinity of the PET radioligand PBR28. J Cereb Blood Flow Metab 2012;32(1):1–5. doi: 10.1038/jcbfm.2011.147 [published Online First: 2011/10/20]

64. Sekine T, Ter Voert EE, Warnock G, et al. Clinical Evaluation of Zero-Echo-Time Attenuation Correction for Brain 18F-FDG PET/MRI: Comparison with Atlas Attenuation Correction. J Nucl Med 2016;57(12):1927–32. doi: 10.2967/jnumed.116.175398 [published Online First: 2016/06/25]

65. Fischl B. FreeSurfer. Neuroimage 2012;62(2):774–81. doi: 10.1016/j.neuroimage.2012.01.021 [published Online First: 2012/01/18]

66. Rousset OG, Ma Y, Evans AC. Correction for partial volume effects in PET: principle and validation. J Nucl Med 1998;39(5):904–11. [published Online First: 1998/05/20]

67. Thomas BA, Cuplov V, Bousse A, et al. PETPVC: a toolbox for performing partial volume correction techniques in positron emission tomography. Phys Med Biol 2016;61(22):7975–93. doi: 10.1088/0031-9155/61/22/7975 [published Online First: 2016/10/27]

68. Hamelin L, Lagarde J, Dorothée G, et al. Early and protective microglial activation in Alzheimer’s disease: a prospective study using 18F-DPA-714 PET imaging. Brain 2016;139(Pt 4):1252–64. doi: 10.1093/brain/aww017 [published Online First: 20160315]

69. Golla SS, Boellaard R, Oikonen V, et al. Quantification of [18F]DPA-714 binding in the human brain: initial studies in healthy controls and Alzheimer’s disease patients. J Cereb Blood Flow Metab 2015;35(5):766–72. doi: 10.1038/jcbfm.2014.261 [published Online First: 2015/02/05]

70. Columba-Cabezas S, Serafini B, Ambrosini E, et al. Induction of macrophage-derived chemokine/CCL22 expression in experimental autoimmune encephalomyelitis and cultured microglia: implications for disease regulation. J Neuroimmunol 2002;130(1-2):10–21. doi: 10.1016/s0165-5728(02)00170-4 [published Online First: 2002/09/13]

71. Harms AS, Thome AD, Yan Z, et al. Peripheral monocyte entry is required for alpha-Synuclein induced inflammation and Neurodegeneration in a model of Parkinson disease. Experimental neurology 2018;300:179–87. doi: 10.1016/j.expneurol.2017.11.010 [published Online First: 2017/11/21]

72. Tuisku J, Plavén-Sigray P, Gaiser EC, et al. Effects of age, BMI and sex on the glial cell marker TSPO — a multicentre [11C]PBR28 HRRT PET study. European Journal of Nuclear Medicine and Molecular Imaging 2019;46(11):2329–38. doi: 10.1007/s00259-019-04403-7

73. Vicente-Rodriguez M, Singh N, Turkheimer F, et al. Resolving the cellular specificity of TSPO imaging in a rat model of peripherally-induced neuroinflammation. Brain Behav Immun 2021;96:154–67. doi: 10.1016/j.bbi.2021.05.025 [published Online First: 2021/05/31]

74. Werry EL, Bright FM, Piguet O, et al. Recent Developments in TSPO PET Imaging as A Biomarker of Neuroinflammation in Neurodegenerative Disorders. Int J Mol Sci 2019;20(13) doi: 10.3390/ijms20133161 [published Online First: 2019/07/03]

75. Wimberley C, Lavisse S, Brulon V, et al. Impact of Endothelial 18-kDa Translocator Protein on the Quantification of (18)F-DPA-714. J Nucl Med 2018;59(2):307–14. doi: 10.2967/jnumed.117.195396 [published Online First: 2017/08/05]

76. Zhou R, Ji B, Kong Y, et al. PET Imaging of Neuroinflammation in Alzheimer’s Disease. Front Immunol 2021;12:739130. doi: 10.3389/fimmu.2021.739130 [published Online First: 2021/10/05]

77. Nutma E, Fancy N, Weinert M, et al. Translocator protein is a marker of activated microglia in rodent models but not human neurodegenerative diseases. bioRxiv 2022;https://doi.org/10.1101/2022.05.11.491453

78. Fang YD, McConathy JE, Yacoubian TA, et al. Image Quantification for TSPO PET with a Novel Image-Derived Input Function Method. Diagnostics (Basel) 2022;12(5) doi: 10.3390/diagnostics12051161 [published Online First: 2022/05/29]

79. Eidson LN, Kannarkat GT, Barnum CJ, et al. Candidate inflammatory biomarkers display unique relationships with alpha-synuclein and correlate with measures of disease severity in subjects with Parkinson’s disease. J Neuroinflammation 2017;14(1):164. doi: 10.1186/s12974-017-0935-1 [published Online First: 2017/08/20]

80. Muñoz-Delgado L, Macías-García D, Jesús S, et al. Peripheral Immune Profile and Neutrophil-to-Lymphocyte Ratio in Parkinson’s Disease. Mov Disord 2021;36(10):2426–30. doi: 10.1002/mds.28685 [published Online First: 2021/06/09]

81. Yu N, Li X, Song W, et al. CD4(+)CD25 (+)CD127 (low/-) T cells: a more specific Treg population in human peripheral blood. Inflammation 2012;35(6):1773–80. doi: 10.1007/s10753-012-9496-8 [published Online First: 2012/07/04]

82. Rocamora-Reverte L, Melzer FL, Wurzner R, et al. The complex role of regulatory T cells in immunity and aging. Front Immunol 2020;11:616949. doi: 10.3389/fimmu.2020.616949 [published Online First: 2021/02/16]

83. Dominguez-Villar M, Hafler DA. Regulatory T cells in autoimmune disease. Nat Immunol 2018;19(7):665–73. doi: 10.1038/s41590-018-0120-4 [published Online First: 2018/06/22]

84. Iglesias JE, Van Leemput K, Bhatt P, et al. Bayesian segmentation of brainstem structures in MRI. Neuroimage 2015;113:184–95.

85. Ewert S, Plettig P, Li N, et al. Toward defining deep brain stimulation targets in MNI space: a subcortical atlas based on multimodal MRI, histology and structural connectivity. Neuroimage 2018;170:271–82.

