## Supplementary Materials, Figure Legends, and Tables for "Brain and systemic inflammation in *de novo* Parkinson’s disease"

**Supplemental Materials.**

**Supplemental Methods.**

Substantia nigra segmentation. The T1 images are processed with FreeSurfer’s ‘Brainstem Substructures’ tool ^84^ which provides the brainstem segmentation of the medulla oblongata, pons, midbrain, and superior cerebellar peduncle. The segmented midbrain is then warped to conform to the midbrain of the DISTAL Atlas^85^ with SPM 8. The transformation of this spatial normalization is then applied to the substantia nigra of the DISTAL Atlas to generate an initial contour of substantia nigra over the PET image space. We then dilated this initial SN contour by three voxels. Over the left side of the dilated initial SN contour, an isocontouring procedure is adopted to extract the top 70 voxels with highest PET intensities to form the left SN contour. The same procedure was repeated with the right SN. The segmented left and right SN contours are then combined to form the final segmentation of the SN VOI.

**Supplemental Figure 1. Participant flow chart.**

**Supplemental Table 1. Demographics of patient cohort**

|  | **PD** | **Control** | ***p*** |
| --- | --- | --- | --- |
|  | **N=58** | **N=62** |  |
| **Age, years** | 66.1 (8.55) | 64.1 (9.10) | 0.180 |
| **Gender:** |  |  | 0.270 |
| Female | 25 (43.1%) | 34 (54.8%) |  |
| Male | 33 (56.9%) | 28 (45.2%) |  |
| **Ethnicity:** |  |  | 0.496 |
| Not Hispanic or Latino | 58 (100%) | 60 (96.8%) |  |
| Not reported | 0 (0.00%) | 2 (3.23%) |  |
| American - Black | 5 (8.62%) | 4 (6.45%) | 0.737 |
| Caucasian | 53 (91.4%) | 58 (93.5%) | 0.737 |
| **Education Level:** |  |  | 0.461 |
| High school graduate | 3 (5.17%) | 8 (12.9%) |  |
| Some college, no degree | 12 (20.7%) | 9 (14.5%) |  |
| Associate degree: occupational, technical, or vocational program | 3 (5.17%) | 4 (6.45%) |  |
| Associate degree: academic program | 3 (5.17%) | 3 (4.84%) |  |
| Bachelors degree | 16 (27.6%) | 18 (29.0%) |  |
| Masters degree | 17 (29.3%) | 14 (22.6%) |  |
| Professional school degree | 2 (3.45%) | 6 (9.68%) |  |
| Doctoral degree | 2 (3.45%) | 0 (0.00%) |  |
| **Employment Status:** |  |  | 0.318 |
| Working now | 18 (31.0%) | 28 (45.2%) |  |
| Retired | 34 (58.6%) | 30 (48.4%) |  |
| Disabled, permanently or temporarily | 1 (1.72%) | 0 (0.00%) |  |
| Keeping house | 2 (3.45%) | 3 (4.84%) |  |
| Other | 3 (5.17%) | 1 (1.61%) |  |

**Supplemental Table 2. Medical history of patient cohort**

|  | **PD** | **Control** | ***p*** |
| --- | --- | --- | --- |
|  | ***N=58*** | ***N=62*** |  |
| **Hypertension** | 30 (51.7%) | 29 (46.8%) | 0.719 |
| **Diabetes mellitus (childhood onset)** | 0 (0%) | 0 (0%) | . |
| **Diabetes mellitus (adult onset)** | 8 (13.8%) | 8 (12.9%) | 1.000 |
| **Myocardial infarction** | 1 (1.72%) | 3 (4.84%) | 0.619 |
| **Congestive heart failure** | 0 (0%) | 0 (0%) | . |
| **Arrhythmia/atrial fibrillation** | 4 (6.90%) | 4 (6.45%) | 1.000 |
| **Hypercholesterolemia** | 31 (53.4%) | 31 (50.0%) | 0.845 |
| **Lung disease** | 1 (1.72%) | 0 (0.00%) | 0.483 |
| **Thyroid disease** | 7 (12.1%) | 10 (16.1%) | 0.707 |
| **Liver disease** | 1 (1.72%) | 2 (3.23%) | 1.000 |
| **Renal disease** | 3 (5.17%) | 1 (1.61%) | 0.352 |
| **Peripheral vascular disease** | 1 (1.72%) | 1 (1.61%) | 1.000 |
| **Stroke or TIA** | 6 (10.3%) | 2 (3.23%) | 0.154 |
| **Seizure** | 1 (1.72%) | 0 (0.00%) | 0.483 |
| **Gaucher disease** | 0 (0%) | 0 (0%) | . |
| **Multiple sclerosis** | 0 (0%) | 0 (0%) | . |
| **Cancer** | 20 (34.5%) | 12 (19.4%) | 0.096 |
| **Other** | 23 (39.7%) | 21 (33.9%) | 0.640 |

**Supplemental Table 3. Family history of patient cohort**

|  | **PD** | **Control** | ***p*** |
| --- | --- | --- | --- |
|  | **N=58** | **N=62** |  |
| **Alzheimer's disease** | 17 (29.3%) | 11 (17.7%) | 0.200 |
| **Amyotrophic lateral sclerosis** | 3 (5.17%) | 1 (1.61%) | 0.352 |
| **Ataxia** | 0 (0%) | 0 (0%) | . |
| **Autism** | 6 (10.3%) | 6 (9.68%) | 1.000 |
| **Bi-polar disorder** | 3 (5.17%) | 4 (6.45%) | 1.000 |
| **Brain aneurysm** | 3 (5.17%) | 8 (12.9%) | 0.250 |
| **Cancer** | 46 (79.3%) | 48 (77.4%) | 0.976 |
| **Dementia** | 23 (39.7%) | 18 (29.0%) | 0.301 |
| **Depression** | 21 (36.2%) | 15 (24.2%) | 0.217 |
| **Diabetes mellitus** | 26 (44.8%) | 25 (40.3%) | 0.753 |
| **Dystonia** | 0 (0%) | 0 (0%) | . |
| **Epilepsy** | 1 (1.72%) | 2 (3.23%) | 1.000 |
| **Heart Disease** | 41 (70.7%) | 40 (64.5%) | 0.599 |
| **Hypertension** | 41 (70.7%) | 37 (59.7%) | 0.284 |
| **Memory loss** | 24 (41.4%) | 20 (32.3%) | 0.397 |
| **Migraines** | 9 (15.5%) | 13 (21.0%) | 0.593 |
| **Multiple sclerosis** | 1 (1.72%) | 3 (4.84%) | 0.619 |
| **Muscle disease** | 1 (1.72%) | 0 (0.00%) | 0.483 |
| **Parkinson's disease** | 11 (19.0%) | 1 (1.61%) | 0.004 |
| **Schizophrenia** | 0 (0.00%) | 1 (1.61%) | 1.000 |
| **Stroke** | 23 (39.7%) | 26 (41.9%) | 0.946 |
| **Suicide or suicide attempt** | 5 (8.62%) | 9 (14.5%) | 0.471 |
| **Tourette syndrome** | 0 (0.00%) | 1 (1.61%) | 1.000 |

**Supplemental Table 4. Vaccination history**

|  | **PD** | **Control** | ***p*** | **N** |
| --- | --- | --- | --- | --- |
|  | **N=58** | **N=62** |  |  |
| **Influenza** | 41 (70.7%) | 39 (62.9%) | 0.477 | 120 |
| **Tetanus, diphtheria, pertussis (Td/Tdap)** | 5 (8.77%) | 0 (0.00%) | 0.023 | 119 |
| **Varicella** | 0 (0%) | 0 (0%) | . | 119 |
| **Zoster** | 7 (12.3%) | 7 (11.3%) | 1.000 | 119 |
| **Measles, mumps, rubella (MMR)** | 0 (0%) | 0 (0%) | . | 119 |
| **Pneumococcal (PCV13 or PPSV23)** | 8 (13.8%) | 4 (6.45%) | 0.301 | 120 |
| **Meningococcal** | 0 (0%) | 0 (0%) | . | 120 |
| **Hepatitis A** | 1 (1.72%) | 1 (1.61%) | 1.000 | 120 |
| **Hepatitis B** | 0 (0%) | 0 (0%) | . | 118 |
| **Haemophilus influenza type B (Hib)** | 0 (0%) | 0 (0%) | . | 120 |
| **COVID-19** | 10 (17.2%) | 25 (40.3%) | 0.010 | 120 |

**Supplemental Table 5. Linear regression analysis of PET imaging.**

| **Brain region** | **Disease**  **(PD vs. HC)** | | **Sex**  **(M vs F)** | | **Genotype**  **(HAB vs MAB)** | |
| --- | --- | --- | --- | --- | --- | --- |
|  | *Estimate* | *p* | *Estimate* | *p* | *Estimate* | *p* |
| **Putamen** | 0.055 | 0.013 | 0.055 | 0.011 | 0.058 | 0.008 |
| **Caudate** | 0.012 | 0.261 | 0.019 | 0.069 | 0.014 | 0.200 |
| **Thalamus** | 0.065 | 0.022 | 0.080 | 0.004 | 0.008 | 0.775 |
| **Substantia nigra** | 0.141 | 0.045 | 0.121 | 0.078 | 0.009 | 0.896 |
| **Brainstem** | 0.018 | 0.354 | 0.018 | 0.347 | -0.054 | 0.007 |
| **Hippocampus** | -0.015 | 0.266 | 0.024 | 0.074 | 0.002 | 0.884 |
| **Frontal cortex** | 0.051 | 0.170 | -0.017 | 0.646 | 0.138 | <.001 |
| **Temporal cortex** | 0.045 | 0.027 | -0.026 | 0.180 | 0.035 | 0.088 |
| **Parietal cortex** | 0.082 | 0.012 | -0.019 | 0.549 | 0.075 | 0.023 |
| **Occipital cortex** | 0.086 | <.001 | -0.028 | 0.256 | 0.034 | 0.180 |

**Supplemental Table 6. Correlation analysis between regional PET binding potential and clinical, cognitive, cytokine/chemokine, and immune cell phenotyping measures in the PD subjects. Separate excel file.**

**Supplemental Table 7. Increased ^18^F-DPA-714 binding potential in PD subjects is predominantly found among high affinity binders.**

|  | **HAB (C/C, N=40)** | | | **MAB (C/T, N=27)** | | |
| --- | --- | --- | --- | --- | --- | --- |
|  | **HC (N=18)** | **PD (N=22)** | ***p*** | **HC (N=19)** | **PD (N=8)** | ***p*** |
| Putamen | 0.07 (0.06) | 0.17 (0.12) | 0.003 | 0.04 (0.05) | 0.05 (0.05) | 0.752 |
| Caudate | 0.01 (0.03) | 0.03 (0.07) | 0.149 | 0.00 (0.01) | 0.00 (0.01) | 0.699 |
| Thalamus | 0.20 (0.14) | 0.31 (0.12) | 0.012 | 0.21 (0.09) | 0.24 (0.08) | 0.405 |
| Substantia nigra | 0.73 (0.33) | 0.92 (0.18) | 0.044 | 0.73 (0.23) | 0.86 (0.42) | 0.434 |
| Brainstem | 0.07 (0.05) | 0.08 (0.09) | 0.768 | 0.11 (0.08) | 0.16 (0.07) | 0.155 |
| Hippocampus | 0.03 (0.08) | 0.03 (0.05) | 0.837 | 0.04 (0.04) | 0.01 (0.02) | 0.052 |
| Frontal cortex | 0.44 (0.14) | 0.49 (0.16) | 0.296 | 0.31 (0.13) | 0.35 (0.13) | 0.436 |
| Temporal cortex | 0.19 (0.08) | 0.23 (0.08) | 0.058 | 0.16 (0.07) | 0.19 (0.09) | 0.490 |
| Parietal cortex | 0.36 (0.15) | 0.46 (0.12) | 0.032 | 0.31 (0.11) | 0.35 (0.10) | 0.312 |
| Occipital cortex | 0.34 (0.11) | 0.43 (0.09) | 0.006 | 0.32 (0.08) | 0.38 (0.11) | 0.242 |
