## Supplementary figures and images for "Brain and systemic inflammation in *de novo* Parkinson’s disease"

### Supplementary Figure 1

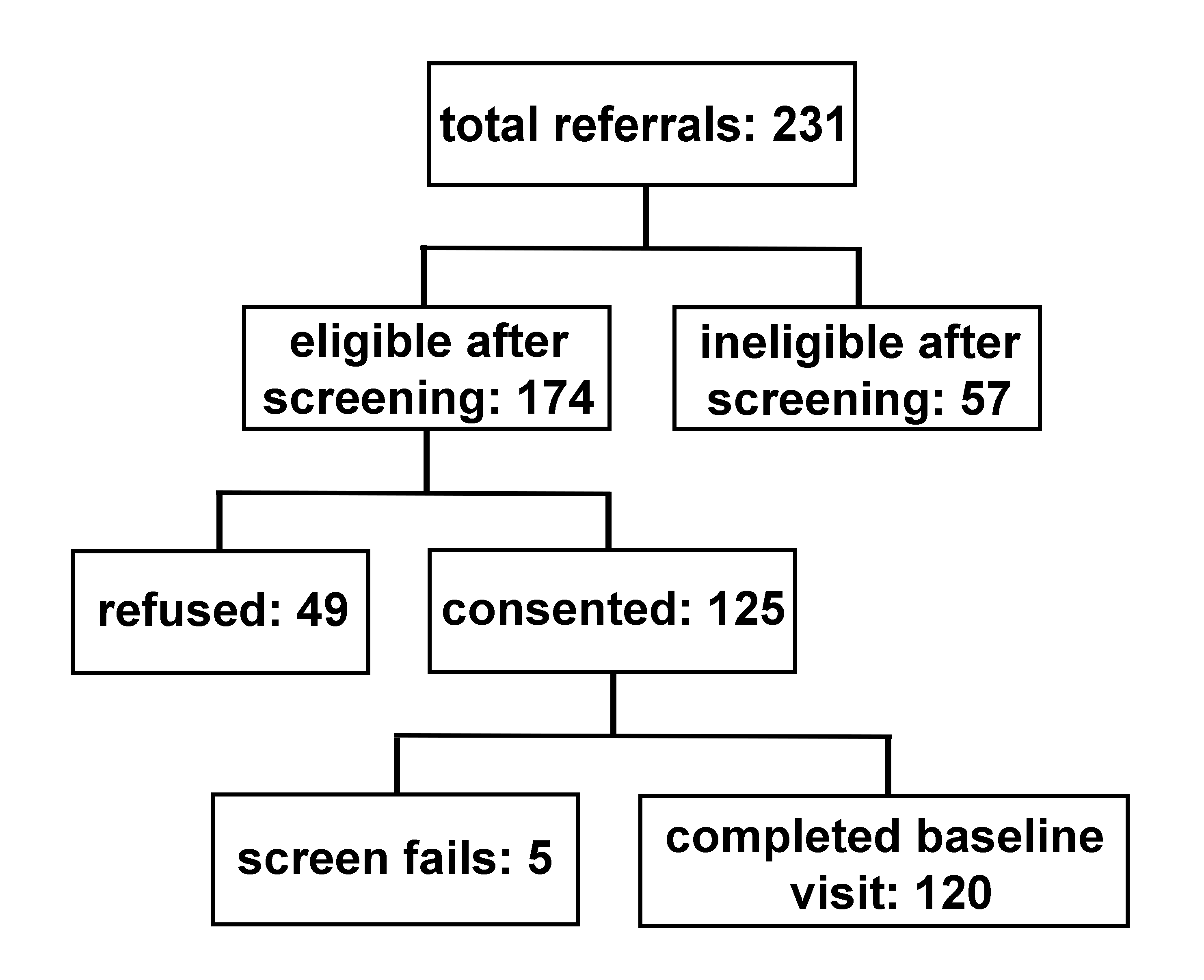
